# Acute post-stroke cognitive test performance predicts one-year functional independence

**DOI:** 10.64898/2025.12.13.25342206

**Authors:** Alan Romanowski, Kathleen Langbehn, Maurizio Corbetta, Daniel Tranel, Aaron D Boes, Mark Bowren

**Author notes:** Corresponding Author: Mark Bowren, Jr., T221 GH, 200 Hawkins Drive, Iowa City, IA 52242. Denotes co-first authors (Alan Romanowski, Kathleen Langbehn). Denotes co-senior authors (Aaron D Boes, Mark Bowren).

## Abstract

**Objective:** General cognitive ability (g), a latent variable derived from cognitive data, can predict life outcomes (e.g., educational attainment and occupational success) among neurologically healthy individuals. The value of g for predicting post-stroke functional outcomes is unknown. We addressed this gap here.

**Method:** We derived g using exploratory structural equation modeling of 15 neuropsychological tests administered to 112 patients with stroke, 69 of whom also had functional outcome data (42 men; mean age = 53.23 years (SD = 10.54)). We used logistic regressions to compare g and individual tests in terms of their ability to predict the Functional Assessment Measure (FAM) and the Functional Independence Measure (FIM; motor and cognitive subscales) at 12 months post-stroke.

**Results:** g was a statistically significant predictor of FAM (*X*^2^ = 6.86, *p* = 0.013) and the cognitive (*X*^2^ = 11.48, *p* = 0.002) but not motor FIM subscale (*X*^2^ = 0.93, *p* = 0.154). Individual tests varied widely in terms of predictive utility (*X*^2^ range: 0.07-21.03), with the most robust predictors being measures of visuospatial functions.

**Conclusions:** Acute measures of cognition, including g, can predict functional independence 12 months after stroke. g and visuospatial ability measures were the most robust predictors.

**Statement of Research Significance:** *Research Question:* General cognitive ability (or, g) is composite score based on scores on performance-based tests of cognition. In neurologically healthy people, g can predict long-term life outcomes. The value of g measured acutely after stroke for predicting functional outcomes one year later is unknown. We evaluated the utility of g for predicting post-stroke functional outcomes.

*Main Findings:* A g factor based on neuropsychological test data collected from acute stroke patients was a statistically significant predictor of cognitive and psychosocial functional outcomes one year later. Lower g after stroke was linked to higher chance of needing functional assistance.

*Study Contributions:* When neuropsychological test data are collected in the acute post-stroke period, it may be possible to derive a g factor score based on those data. Our findings suggest that this g factor score could help clinicians tailor treatment strategies to a patient’s expected long-term functional outcomes.

## Introduction

Long-term functional disability is prevalent among stroke survivors (Sennfält et al., 2019; Veerbeek et al., 2011). The ability to identify, in the acute setting, patients at an elevated risk for post-stroke disability could help clinicians optimize treatment strategies and healthcare plans. Performance-based behavioral measures are often administered for this purpose. The most widely used such measure is the NIH Stroke Scale, which can predict many post-stroke functional outcomes (Bustamante et al., 2016) but has limited coverage of higher-order cognitive abilities. A variety of cognitive and neuropsychological tests have been shown to predict post-stroke functional outcomes (Nys et al., 2005), and are capable of adding incremental predictive value beyond the NIH Stroke Scale (Sharma et al., 2020; Wagle et al., 2011). However, the potential utility of cognitive composite measures, which are typically more psychometrically reliable than individual tests from which they were derived, has received comparatively less attention.

General cognitive ability (g) is a latent variable that is statistically derived from the shared variance across diverse cognitive test performances (Spearman, 1904). Prior studies have shown that g can predict outcomes in neurologically healthy populations, including academic achievement and job performance (Gottfredson, 2003; Spinath et al., 2006). Because variance attributable to g is stable across different test batteries, g can be derived from any diverse battery of cognitive tests (Johnson et al., 2004, 2008), including typical neuropsychological test batteries. Thus, acute g could be readily calculated from any existing clinical protocol for neuropsychological testing, and could serve as flexible, data-driven supplement to other outcome prediction measures. However, the value of g for predicting chronic functional outcomes among patients with stroke has not been studied.

Here, we used exploratory structural equation modeling to calculate latent g using a hierarchical model based on acute post-stroke neuropsychological test data in 112 patients with first-ever stroke. Then, for a subset of participants with functional outcome data, we used g to predict three widely used functional outcome scores that measure real-world functioning across cognitive, psychosocial, and motor domains. We hypothesized that acute g would predict cognitive and psychosocial outcomes, but not motor outcomes measured one year after stroke.

## Method

### Participants

Patients with new-onset ischemic or hemorrhagic stroke were identified from a prospectively enrolled sample collected at Washington University in St. Louis (WashU) which has been described previously (Corbetta et al., 2015). Inclusion criteria included: evidence of neurological deficit, wakefulness and alertness, and ability to participate in research. Exclusion criteria included previous stroke and significant history of neurological, psychiatric or medical conditions. Participants provided informed consent, and the study and analyses were approved by the Institutional Review Board at WashU. Analyses were also approved by the Institutional Review Board at the University of Iowa.

Modeling of g (described below) was performed using neuropsychological data from all participants who completed such testing within 14 days (median = 12 days; n = 112). Individual neuropsychological tests and g scores were associated with functional measures for a subset of patients who also had functional outcomes measured 12 months post-stroke. This subsample was used for our primary analyses (i.e., outcome predictions; n = 69). This sample comprised 42 men and 27 women, with a mean age of 53.23 years (SD = 10.54), and mean education of 13.68 years (SD = 2.60). There were 46 African American individuals and 23 White individuals. One participant was Hispanic and the remaining 68 were non-Hispanic. There were 63 right-handed and 6 left-handed individuals. There were 33 participants with left hemisphere stroke and 36 with right hemisphere stroke.

### Functional Outcomes

The Functional Independence Measure (FIM) was developed as a sensitive and comprehensive measure of disability, with individual items focusing on specific activities of daily living (e.g., toileting, grooming, walking, problem solving). Items are scored based on how much assistance is required to perform them (Keith et al., 1987). This assessment is administered by a trained evaluator, with scoring based on observation, interview, and information from medical records. Items are scored on a 7-point scale based on the amount of dependence required to perform tasks. An item score of 1 represents total dependence, and 7 represents total independence. The 18 items are separated into two subscales: Motor (13 items) and Cognitive (5 items).

The Functional Assessment Measure (FAM) is a 12-item measure of additional functional outcomes. It was developed as an adjunct to the FIM, and it is designed to address major functional areas which are less emphasized in the FIM, many of which are broadly within the psychosocial domain (e.g., employability, community access, adjustment to limitations), with comparatively fewer items addressing other areas, such as swallowing and car transfer. Administration and scoring are identical to FIM. The FAM is not intended as a standalone measure, and in clinical practice, is administered along with the FIM (Hall et al., 1993).

The FAM total score, and FIM cognitive and motor subscales were the dependent variables for our analyses. The distributions of all three of the functional outcome measures were skewed, with most participants rated as being completely independent across all items (i.e., maximum scores on each scale). Thus, for each measure, we binarized scores into one of two conditions: Independent (no help needed; maximum score on the measure: score of 84 for the FAM, score of 35 for the FIM cognitive subscale, score of 91 for the FIM motor subscale), or Assistance Needed (any degree of help needed; scores less than the maximum).

### Neuropsychological Tests

The final set of behavioral measures comprised 15 neuropsychological tests: the Boston Diagnostic Aphasia Exam (BDAE subtests: Boston Naming Test (short form), Word Comprehension, Commands, Complex Ideational Material, Oral Reading of Sentences, Comprehension of Oral Reading of Sentences); Animal Naming Test; Brief Visuospatial Memory Test (BVMT; subtests: immediate recall, delayed recall, and recognition discrimination index); Hopkins Verbal Learning Test Revised (HVLT; subtests: immediate recall, delayed recall, recognition discrimination index); the Spatial Span subtests from the Wechsler Memory Scale Third Edition (separate scores for forward and backward spans). There were three tests of hemispatial inattention/neglect which were not used in our analyses because of heavy skew of their distribution resulted in lack of model convergence in both the exploratory structural equation modeling and logistic regressions. Those tests were: Mesulam Cancellation Test (total misses), Posner Cueing Task (average accuracy), and the Star Cancellation subtest from the Behavioral Inattention Test (total misses). We used normatively adjusted scores when available as described in the original study for this cohort (e.g., based on test manuals; Corbetta et al., 2015).

### Calculation of g

We derived g from the neuropsychological tests described above using a hierarchical model of cognition within an exploratory structural equation modeling framework. This analysis was performed using the lavaan package in R version 4.4.1. In exploratory structural equation modeling, some factor loadings (in this case, loadings of tests onto the first-order latent factors) are calculated in a data-driven manner, and the others (in our case, the loadings of the latent factors onto the g factor, as well as test covariance between Spatial Span forward and backward) are specified in the model. Notably, exploratory structural equation modeling allows for cross-loadings onto the first-order factors, which is useful because any given neuropsychological test rarely measures one and only one cognitive function. We used parallel analysis to determine the number of first-order latent factors to model. A parallel analysis suggests the number of factors to model by comparing eigenvalues calculated from the true dataset to eigenvalues calculated from randomly generated data matrices of the same size. Structural equation modeling was performed using the robust maximum likelihood factoring method (i.e., estimator was set to “MLR” in lavaan). All individual neuropsychological tests were centered and scaled prior to entry into the model. Although the focus of this study was on g, we also used the first-order latent factors as predictors as they were readily available. We evaluated the fit of our exploratory structural equation model using the following global fit indices: Comparative Fit Index (CFI), Tucker-Lewis Index (TLI), Root Mean Square Error of Approximation (RMSEA), and Mean Squared Residual (RMSR). Typical cutoffs for acceptable model fit are: 0.90-0.95 or above for CFI and TLI, less than 0.08 for RMSEA, and less than 0.05 for SRMR. We reported the fully standardized factor loadings.

### Logistic Regressions

We used the glm function in R to perform logistic regressions. In each model, we predicted the binarized versions of the FAM, FIM cognitive subscale, and FIM motor subscale using one of the neuropsychological tests or latent factor scores from the exploratory structural equation model (19 total predictors). We built separate univariate models (i.e., models with one predictor of interest) for each combination of dependent variable and predictor. The final logistic regression model results were adjusted for multiple comparisons using False Discovery Rate (FDR) with a threshold rate of 5% for error rate control. To compare the relative predictive value of the various models, we used the *X*^2^ metric, where larger values indicate greater effect size. We did not compare models based on Odds Ratio because this metric is dependent upon the scale of the underlying predictor variables, which was not uniform across neuropsychological tests.

## Results

### Exploratory Factor Analysis

Parallel analysis supported three first level latent factors. The model converged normally. Model fit was in the acceptable range for all fit measures: CFI = 0.99, TLI = 0.98, RMSEA = 0.04, SRMR = 0.03. Based on the tests loading onto each factor, the first-order factors broadly corresponded to visuospatial ability/memory, verbal memory, and language. The factor loadings from the EFA are shown in Figure 1. Interestingly, the visuospatial ability/memory factor had by far the strongest loading onto the g factor.

**Figure 1.**
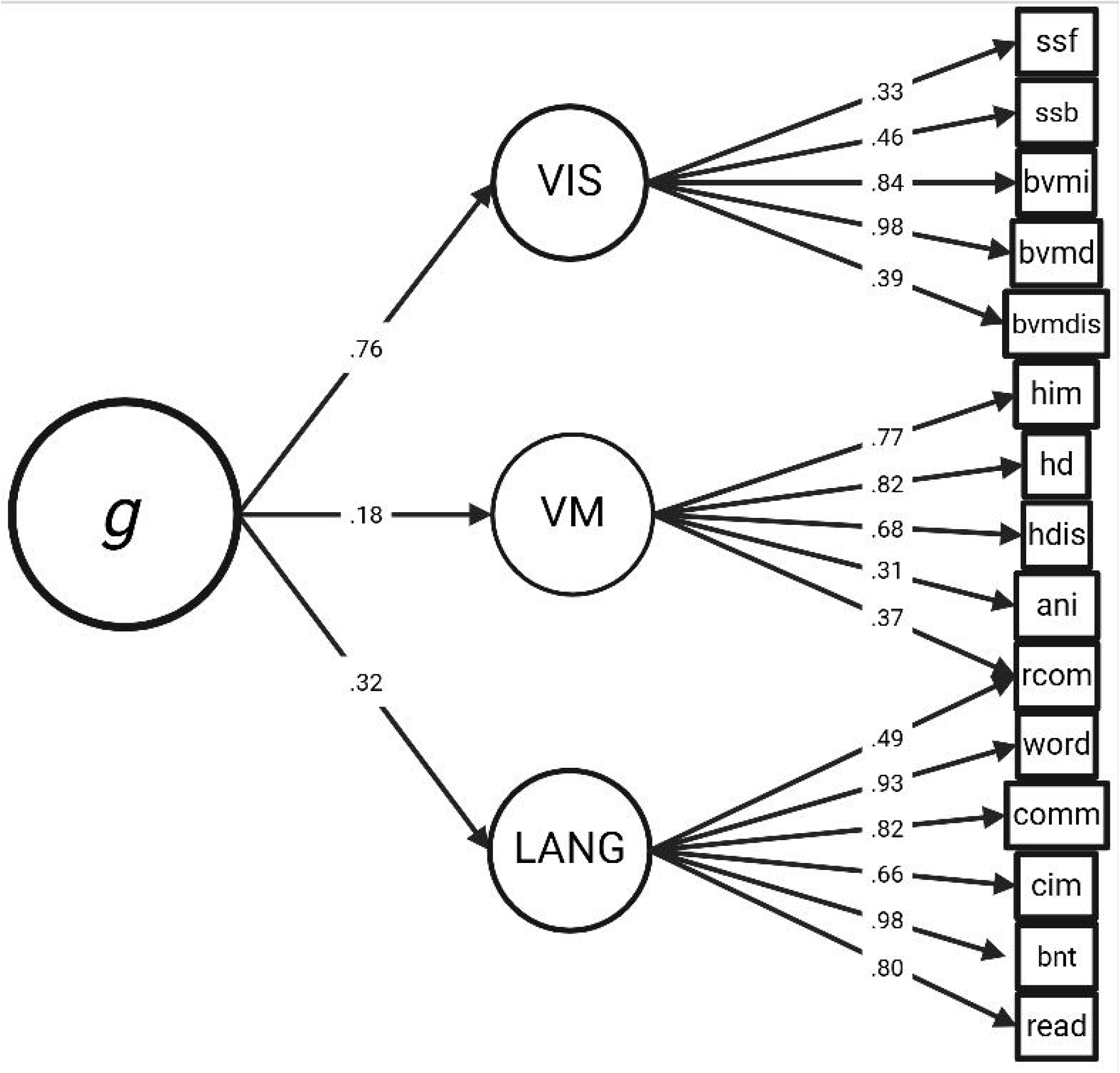
Exploratory Structural Equation Model for Deriving g. The R package lavaan was used to create an exploratory structural equation model based on the neuropsychological test data. The first-order latent factors were derived in a data-driven manner, and each test was allowed to load onto each factor. For clarity, only factor loadings greater than 0.30 are displayed. The first-order factors were labelled as visuospatial ability/memory (VIS), verbal memory (VM), and language (LANG) based on the pattern of loadings associated with each factor. Each first-order factor then loaded onto a single domain-general cognitive factor (g). ssf = Spatial Span forward; ssb = Spatial Span backward; bvmi = BVMT Immediate Recall; bvmd = BVMT Delayed Recall; bvmdis = BVMT Recognition Discrimination; him = HVLT Immediate Recall; hd = HVLT Delayed Recall; hdis = HVLT Discrimination Index; ani = Animal Naming Test; rcom = Comprehension of Oral Reading of Sentence; word = Word Comprehension; comm = Commands; cim = Complex Ideational Materia; bnt = Boston Naming Test; read = Oral Comprehension of Sentences.

### Logistic Regressions

Details of statistical results from the logistic regressions are presented in Table 1. For brevity, Table 1 focuses on all latent factor-related results (i.e., g and the other factors from the exploratory structural equation model), and all model results for the individual tests that survived FDR correction for multiple comparisons. g was a statistically significant predictor of both FAM (*X*^2^ = 6.86; FDR q = 0.041) and the cognitive subscale of the FIM (*X*^2^ = 11.48; FDR q = 0.016), but not the motor subscale of the FIM (*X*^2^ = 2.08; FDR q = 0.244). The same was true for the visuospatial ability/memory factor (FAM model *X*^2^ = 7.77, FDR q = 0.039; FIM cognitive subscale model *X*^2^ = 14.20, FDR q = 0.011; FIM motor subscale model *X*^2^ = 1.93, FDR q = 0.248), and the language factor (FAM model *X*^2^ = 7.77, FDR q = 0.041; FIM cognitive subscale model *X*^2^ = 15.26, FDR q = 0.011; FIM motor subscale model *X*^2^ = 1.75, FDR q = 0.270).

**Table 1.** Functional outcome measures at 12-months predicted by acute factor scores and individual neuropsychological tests.

| *Predictors* | *Odds Ratios* | *Odds Ratio 95% CI* | *Z-stat* | *Chi-squared* | *p-value* | *FDR q-value* |
| --- | --- | --- | --- | --- | --- | --- |
| **FAM** |  |  |  |  |  |  |
| g | 2.29 | 2.49 – 4.56 | 2.49 | 6.86 | **0.013** | **0.041** |
| Visuospatial Ability/Memory Factor | 1.59 | 1.14 – 2.28 | 2.64 | 7.77 | **0.008** | **0.039** |
| Verbal Memory Factor | 1.63 | 0.90 – 3.07 | 1.58 | 2.61 | 0.114 | 0.191 |
| Language Factor | 3.15 | 1.38 – 8.37 | 2.52 | 7.77 | **0.012** | **0.041** |
| Spatial Span Forward | 1.28 | 1.07 – 1.57 | 2.59 | 7.49 | **0.010** | **0.041** |
| Spatial Span Backwards | 1.49 | 1.24 – 1.86 | 3.86 | 21.03 | **<0.001** | **0.006** |
| BVMT Delay | 1.03 | 1.01 – 1.06 | 2.66 | 7.86 | **0.008** | **0.041** |
| **FIM Cognitive Subscale** |  |  |  |  |  |  |
| g | 3.76 | 1.70 – 9.50 | 3.06 | 11.48 | **0.002** | **0.016** |
| Visuospatial Ability/Memory Factor | 2.22 | 1.43 – 3.76 | 3.30 | 14.20 | **0.001** | **0.011** |
| Verbal Memory Factor | 2.70 | 1.29 – 6.28 | 2.49 | 7.08 | **0.013** | **0.041** |
| Language Factor | 6.03 | 2.32 – 19.58 | 3.36 | 15.26 | **<0.001** | **0.011** |
| Word Comprehension | 1.06 | 1.03 – 1.12 | 2.96 | 12.64 | **0.003** | **0.017** |
| Complex Ideational Material | 1.04 | 1.02 – 1.07 | 3.16 | 11.98 | **0.002** | **0.013** |
| Reading Comprehension | 1.03 | 1.01 – 1.05 | 2.99 | 9.91 | **0.003** | **0.017** |
| Animal Naming | 1.04 | 1.01 – 1.07 | 2.46 | 8.77 | **0.014** | **0.042** |
| HVLT Immediate Recall | 1.05 | 1.01 – 1.09 | 2.52 | 6.87 | **0.012** | **0.041** |
| HVLT Recognition Discrimination | 1.04 | 1.01 – 1.07 | 2.56 | 6.95 | **0.010** | **0.041** |
| BVMT Immediate Recall | 1.07 | 1.03 – 1.11 | 3.34 | 16.01 | **<0.001** | **0.011** |
| BVMT Delayed Recall | 1.06 | 1.03 – 1.10 | 3.32 | 14.42 | **<0.001** | **0.011** |
| **FIM Motor Subscale** |  |  |  |  |  |  |
| g | 1.55 | 0.86 – 2.91 | 1.42 | 2.08 | 0.154 | 0.244 |
| Visuospatial<br>Ability/Memory Factor | 1.25 | 0.91 – 1.73 | 1.37 | 1.93 | 0.170 | 0.248 |
| Verbal Memory Factor | 1.06 | 0.59 – 1.91 | 0.19 | 0.03 | 0.853 | 0.853 |
| Language Factor | 1.64 | 0.79 – 3.58 | 1.30 | 1.75 | 0.195 | 0.270 |
| Spatial Span Backward | 1.33 | 1.13 – 1.60 | 3.21 | 12.55 | **0.001** | **0.013** |

Results shown are for individual logistic regression models. FDR adjusted q-values shown are corrected for multiple comparisons across all model combinations of predictors and dependent variables. Z-statistic indicates the signed square root of the chi-square statistic from logistic regression models.

Regarding individual tests, FAM was statistically significantly predicted by Spatial Span Forwards and Backwards, and BVMT Delayed Recall. The FIM cognitive subscale was significantly predicted by Word Comprehension, Complex Ideational Material, Comprehension of Oral Reading of Sentences, HVLT Immediate Recall, HVLT Recognition Discrimination, BVMT Immediate Recall, BVMT Delayed Recall, and the Animal Naming Test. Lastly, the only statistically significant predictor of the FIM motor subscale was Spatial Span Backward. Compared to the latent factors described in the paragraph above, Spatial Span Forward and Backward, and BVMT Delayed Recall had higher *X*^2^ values for prediction of the FAM. The same was true for BVMT Immediate Recall for predicting the FIM cognitive subscale (*X*^2^ = 16.01). Thus, measures of visuospatial ability/memory were, in addition to the factor scores from the exploratory structural equation model, generally robust predictors of different types of functional outcome measures.

## Discussion

A latent g factor derived from acutely administered neuropsychological tests was a statistically significant predictor of functional outcomes measured 12 months post-stroke. This g factor was most strongly predictive of cognitive functional outcomes as measured by the FIM, though it was also significantly predictive of outcomes as measured by the FAM, which largely measures psychosocial aspects of functioning. As predicted, g was not statistically significantly associated with 12-month motor outcomes measured by the FIM. Other latent cognitive factors reflecting visuospatial ability/memory and language were also statistically significant predictors of cognitive and psychosocial outcomes. In sum, our findings support the notion that latent variables derived from acute neuropsychological data can be used to predict long-term cognitive and psychosocial functional outcomes after stroke.

Our findings support the use of neuropsychological tests and associated latent variables for predicting cognitive and psychosocial, but not motor, functional outcomes after stroke. All three latent cognitive variables and 8 of the 15 individual neuropsychological tests were statistically significant predictors of cognitive functional outcomes, but only one test was a statistically significant predictor of chronic motor outcomes. Due to their generally low utility for predicting motor outcomes, it will remain important to combine neuropsychological measures with tools designed to predict functional motor outcomes.

Among the individual tests, those that were most robustly predictive tapped visuospatial processing, such as the Spatial Span test. These findings are consistent with the logistic regression results based on the latent factors, as the visuospatial ability/memory factor was associated with the best predictive metrics. Notably, this factor was also the first-order factor most strongly linked to g in this sample. The Spatial Span measures may have particularly high predictive value because they measure working memory, which is tightly linked to g and is thought to play a role in many forms of higher-order cognition (Bowren et al., 2020; Shipstead & Yonehiro, 2016). Also, visuospatial measures are often sensitive to hemispatial inattention/neglect, which has been linked to poor functional outcomes in prior work (Di Monaco et al., 2011). Regardless of the reason for the predictive utility of the visuospatial measures used in this study, our findings suggest that prioritizing visuospatial tests during acute post-stroke neuropsychological exams will, on balance, yield prognostically valuable information. However, we emphasize that the most valuable test(s) for predicting long-term outcomes likely varies from patient to patient. The predictive value of tests of language may, for example, be higher for patients with acute aphasia. Thus, a hypothesis-driven approach remains key to planning time-limited neuropsychological exams in the setting of acute stroke.

This study is limited by the scope of the available behavioral measures. Many other neuropsychological and cognitive tests that could prove to be as or more valuable for functional outcome predictions than those used here. As is often the case, our study is also limited by the skewed distribution of functional outcome scores in our sample (Turner-Stokes & Siegert, 2013). This limited our ability to make more precise claims about the level of disability that can be predicted. We are limited to claiming that the measures in this study can predict whether a patient will go on to need at least some functional assistance 12 months after stroke. Lastly, future studies would be needed to evaluate the additive value of cognitive, psychosocial, and lesion factors for predicting post-stroke outcomes.

In summary, the present study provides empirical evidence for the predictive utility of neuropsychological tests and associated latent variables for predicting 12-month post-stroke outcomes. These findings could lead to improvements in techniques for prognostication based on acute behavioral data.

## Data Availability

All data produced in the present study are available upon reasonable request to the authors.

## Acknowledgements

Alan Romanowski: COI: None

Kathleen Langbehn: COI: None

Maurizio Corbetta: COI: None

Daniel Tranel: COI: None

Aaron D. Boes: COI: None

Mark Bowren Jr: COI: None

This work was supported by the National Institute of General Medical Sciences (M.B, A.B., D.T, T32GM108540), (M.C., T32 GM007200); the National Institutes of Mental Health (A.R., K.L., M.B., A.B., D.T., 1 P50 MH094258), (A.R., K.L., M.B., A.B., D.T., 1 R21 MH120441-01), (A.R., K.L., M.B., A.B., D.T., 2 T32MH019113-29A1), (M.C., F30 MH099877); the National Institute of Child Health and Human Development (M.C., R01 HD068290), (M.C., R01 HD061117-05), (M.C., R01 HD068290-03); the Kiwanis Foundation (A.R., K.L., M.B., A.B., D.T.); FC-Neuro University of Padua (M.C.); the National Institute of Neurological Disease and Stroke (A.R., K.L., M.B., A.B., D.T., 1 R01 NS114405-01), (A.R., K.L., M.B., A.B., D.T., NS095741); FLAG-ERA JTC (M.C.); Neuro-DiP: Progetto Dipartimenti di Eccellenza Italian Ministry of Research (MIUR) (M.C.); CARIPARO Foundation Padova (M.C.); and the University of Iowa Roy J. and Lucille A. Carver College of Medicine (A.R., K.L., M.B., A.B., D.T.). This work was conducted on an MRI instrument funded by 1S10RR028821-01 (M.C.).

## Notes

### Competing Interest Statement

The authors have declared no competing interest.

### Author Declarations

IRB of Washington University in St. Louis gave ethical approval for this work. IRB of University of Iowa gave ethical approval for this work.

